# Bayesian hybrid statistical and machine learning models for dengue forecasting in Bangladesh: Temporal and spatial analysis for an early warning system

**DOI:** 10.1101/2025.09.14.25335716

**Authors:** Shamima Hossain, Mahia Mehrun Safa, Nahid Farzana Juthi, Nowshin Tasnia

## Abstract

Dengue remains a major public health concern in Bangladesh, yet reliable forecasting models that integrate climatic and demographic drivers are limited. Developing an early warning system (EWS) capable of anticipating outbreaks is critical for effective prevention and control. We analysed hospital-based dengue surveillance data covering admissions from January 2000 to August 2025 alongside climatic (temperature, rainfall, humidity) and demographic (population density, proportion of urban population) covariates. A suite of Bayesian statistical mixture and machine learning hybrid models, including SARIMA–Poisson, SARIMA–negative binomial (NB), SARIMA–SVM, SARIMA–LSTM, and SARIMA–XGBoost, were evaluated. Model performance was assessed using Leave-One-Out Information Criterion (looic), Root Mean Square Error (RMSE), Mean Absolute Error (MAE), Continuous Ranked Probability Score (CRPS), and coverage probability (CVG). Sensitivity and specificity were also computed to assess early warning performance. Spatial dependence was examined using Global Moran’s I and Local Moran’s I (LISA) cluster maps by using district level monthly dengue hospitalize cases data for 2019, and from 2022 to 2024. Rainfall and portion of urban population emerged as significant drivers of dengue incidence, while temperature, humidity, and population density were less influential. Global Moran’s I indicated no significant spatial autocorrelation at the district level; however, LISA maps identified localised hotspots. Among the candidate models, the Bayesian SARIMA–XGBoost hybrid achieved the best predictive performance, with the lowest Continuous Ranked Probability Score (CRPS) and the highest coverage probability (CVG), providing the most balanced sensitivity–specificity trade-off. Forecasts for January to August 2025 accurately reproduced seasonal dynamics, predicting a sharp rise during the monsoon, with peak incidence in July. Although magnitudes were overestimated, outbreak timing was well captured. The Bayesian SARIMA–XGBoost hybrid model offers a robust framework for probabilistic dengue forecasting in Bangladesh. By linking upstream surveillance data to forecast production, this study demonstrates the potential for a fully implemented early warning system (EWS) to strengthen outbreak preparedness. Future work should incorporate finer spatial resolution, real-time climate forecasts, and entomological indicators to enhance operational deployment.

## Introduction

Dengue fever remains a rapidly escalating global threat: nearly half the world’s population is at risk, and recent seasons have produced unprecedented case counts and geographic spread [1–3]. Bangladesh exemplifies this shift. In 2023 the country experienced its deadliest epidemic on record, with 321,179 cases and 1,705 deaths, underscoring the need for operationally useful forecasts and targeted risk mapping [4].

Within Bangladesh, dengue has evolved from a Dhaka-centric problem into a broader, district-level challenge. Recent work has identified changing hotspots, strong seasonality, and spatial clustering across the country, calling for locality-specific risk assessment rather than single-city strategies [5–7]. Parallel studies in the region show that method choice for spatiotemporal hotspot detection materially affects decisions on where to intervene [8].

Classical time-series models (e.g., ARIMA/SARIMA, ETS/TBATS) remain useful baselines for monthly surveillance data and can capture regular seasonality; yet they often struggle with nonlinearity, regime shifts, over-dispersion, and frequent zeros that characterize dengue counts. In Bangladesh specifically, time-series analyses signal strong climate–incidence associations and predict multiple-year oscillations, but with limited performance during abrupt epidemic transitions [9–11].

Consequently, machine-learning (ML) pipelines have gained traction. For Bangladesh, a Scientific Reports study integrated 13 meteorological predictors and tailored feature engineering to deliver high monthly accuracy and realistic seasonal peaks [12]. Interpretable tree-based systems (e.g., Light GBM with SHAP) have been adapted into early-warning tools using climate, landscape, and sociodemographic covariates [13], and explainable ML approaches (e.g., XGBoost) have been proposed to enhance outbreak prediction transparency [14]. Comparable advances in Singapore and Brazil demonstrate that modern ML (XGBoost, LSTM, ensembles) improves short-horizon forecasts and can incorporate spatial lags from neighbouring areas to capture diffusion [15–17].

Data engineering further boosts performance. In Bangladesh, stochastic Bayesian downscaling of surveillance series followed by ML has improved temporal resolution and accuracy for operational forecasting [18]. At the entomological layer, spatial downscaling has been used to refine vector-risk maps, supporting finer-scale prevention [19].

Hybrid frameworks that marry linear structure with nonlinear learners are particularly promising. The ARIMA–NNAR family explicitly models the linear component and learns the residual nonlinearity, while ARIMA–ARNN variants admit exogenous climate drivers for scenario exploration; recent seasonal-adjusted hybrid ML algorithms similarly target complex seasonal patterns at scale [20–22].

Equally important is distributional realism for count data. Bangladesh dengue series are commonly over-dispersed and zero-inflated, motivating mixture and hurdle models. City-level analyses in Dhaka using count-regression frameworks highlight non-Gaussian noise and covariate effects [23]; long-term predictors discovered via data-mining emphasize multi-factor drivers [24]. Mixture approaches including Zero inflated negative binomial (ZINB) and related formulations have been recommended in comparative studies and applied in dengue prediction tasks, including hybrid machine learning (ML) and zero-inflated two-stage workflows for presence or abundance [25–28]. Recent statistical work extends the endemic–epidemic (HHH) family with zero-inflation to handle sparse surveillance while preserving interpretability for public-health use [26,29].

Because dengue risk is inherently spatial, policy-ready models must quantify neighbourhood diffusion and spatial random effects. The endemic–epidemic (HHH) framework decomposes endemic seasonality from epidemic autoregression and spatial spillover [30], while Bayesian hierarchical models via INLA with CAR/BYM priors enable efficient district-level mapping with uncertainty quantification [31–33]. Applications across Bangladesh and Southeast Asia (e.g., Laos, Indonesia) confirm the value of INLA-based spatiotemporal models that integrate climate, environment, and demography to produce interpretable risk surfaces for targeting [32–33].

Finally, for real-time decision support, nowcasting to correct reporting delays can stabilize short-term situational awareness. Bayesian nowcasting frameworks originally demonstrated for dengue in Bangkok have proven effective and transferable, and general nowcasting toolkits (e.g., NobBS) formalize uncertainty under varying delay structures [34,35].

Within this comprehensive epidemiological and methodological landscape, our study benchmarks five SARIMA–Bayesian mixture models—combining seasonal autoregressive structure with Bayesian inference and alternative learners: Poisson regression, negative-binomial regression, XGBoost, SVM, and LSTM to identify the optimal approach for forecasting monthly dengue cases across Bangladesh. Specifically, our objectives are to: (i) benchmark predictive accuracy across these five models; (ii) evaluate model performance using the leave-one-out information criterion (LOOIC); (iii) generate short-term uncertainty-quantified forecasts; (iv) produce district-level risk maps to support targeted vector-control efforts; and (v) identify which climate and demographic variables significantly affect dengue cases. By directly addressing nonlinearity, over-dispersion, zero-inflation, and spatial dependence while leveraging rich climate and demographic data, this work aims to respond effectively to Bangladesh’s evolving dengue epidemiology and contribute actionable insights for disease control [36–42].

## Materials and methods

### Data source

#### Data on Dengue fever cases

The Directorate General of Health Services (DGHS) receives reports of suspected, probable, and confirmed dengue cases that have been seen in medical facilities around the nation. Suspected cases are those who have acute febrile illness with or without nonspecific symptoms. In contrast, likely cases are those with an acute febrile illness with a serological diagnosis. The confirmed cases should have an acute febrile illness with a positive dengue NS1 antigen or PCR test. Details of dengue case definition and management are available from the DGHS. Daily reports of confirmed dengue cases are compiled and circulated by the DGHS’s communicable disease control (CDC) unit. Monthly dengue cases were collected from the DGHS from January 2000 to August 2025.

#### Meteorological Data

The Bangladesh Meteorological Department (BMD) operates 35 weather stations across the country, which record daily temperature (°C), rainfall (mm), and relative humidity (%). For this study, we aggregated daily station data into monthly averages (for temperature and humidity) and monthly totals (for rainfall) for each year in the study period.

#### Demographic Data

National population data from 2000 onward were collected from multiple rounds of BBS population censuses (2001, 2011, and 2022) [43–45]. Intercensal years were estimated using official BBS growth rates, complemented by World Bank demographic indicators [46]. Population density was calculated annually as the ratio of total national population to total land area. The percentage of the urban population was derived from BBS census reports [43–45] and interpolated for intercensal years using linear trends. For 2023 and 2024, national population density and urban share were projected from 2022 values using a uniform annual growth rate of 1.1% [45,46].

### Exploratory data analysis

#### Basic Characteristics

A total of 666,639 dengue fever cases and 3,139 dengue-related deaths were reported in Bangladesh between January 2000 and December 2024. The annual number of cases varied considerably, ranging from 375 cases in 2014 to 321,179 cases in 2023. Dengue mortality showed similar fluctuations, with no recorded deaths in several years between 2007 and 2014, rising to a peak of 1,705 deaths in 2023. The epidemics of 2019 (101,354 cases; 170 deaths) and 2023 (321,179 cases; 1,705 deaths) represent the severe outbreaks during the study period. Importantly, in 2024, the number of dengue-related deaths reached 575, the second-highest figure on record, emphasising heightened transmission intensity and increased mortality risk. This underlines the urgent need for effective public health interventions and robust disease management strategies. Notably, the 2023 epidemic alone accounted for more than 180 times the number of cases reported in 2014 and contributed to over 54% of all reported dengue deaths nationwide. These findings demonstrate a highly skewed temporal distribution, with rare but extremely intense outbreak years dominating the overall burden (S1 Fig).

#### Regional variations

To assess geographic heterogeneity in dengue incidence, we first examined division-level distributions of total reported cases in Bangladesh for the period 2019–2024. The choropleth highlights clear regional differences, with the highest dengue burden concentrated in Dhaka and Chittagong divisions, while peripheral divisions such as Rangpur and Sylhet reported relatively lower-case counts (S2 Fig). To explore these patterns in greater detail, district-level monthly distributions were mapped for 2022, 2023, and 2024. These visualisations reveal persistent hotspots where dengue cases remain consistently high, particularly in central and southeastern districts, whereas some northern and coastal districts consistently experience very low incidence (S3-S5 Figs). Further evidence of regional variation was obtained from the distribution of annual dengue cases across districts from 2000–2024 (log_10_ scale) (S6 Fig). The boxplot demonstrates wide variation, with certain districts exhibiting consistently elevated incidence, while others remain at the lower end of the distribution. These findings collectively underscore substantial spatial heterogeneity in dengue transmission across Bangladesh, highlighting the presence of persistent high-risk districts that may warrant targeted surveillance and control interventions.

#### Spatial Dependency

Global Moran’s I values for 2019 to 2024 were close to zero (Table 1), indicating no strong nationwide spatial clustering of dengue incidence. In 2019 (I = –0.0096, p = 0.2810) and 2022 (I = –0.0118, p = 0.2640), the statistics were slightly negative, while in 2023 (I = 0.0192, p = 0.0760) and 2024 (I = 0.0319, p = 0.0530), values were weakly positive but remained non-significant.

**Table 1.**
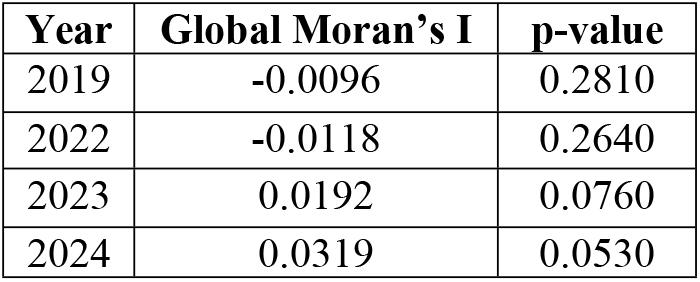
Global Moran’s I statistics for district-level dengue incidence in Bangladesh, 2019– 2024.

| Year | Global Moran’s I | p-value |
| --- | --- | --- |
| 2019 | -0.0096 | 0.2810 |
| 2022 | -0.0118 | 0.2640 |
| 2023 | 0.0192 | 0.0760 |
| 2024 | 0.0319 | 0.0530 |

Global Moran’s I values provide a measure of overall spatial autocorrelation in dengue incidence across districts. The results indicate no statistically significant global clustering at the 5% level during the study period, suggesting that dengue incidence was not uniformly clustered at the national scale.

In contrast, Local Moran’s I (LISA) revealed more informative district-level patterns (Fig 1). In 2019 and 2022, widespread positive autocorrelation (high–high clusters in red) suggested spatial clustering, although Dhaka occasionally appeared as a high-value outlier. By 2023 and 2024, most districts exhibited weak autocorrelation (blue), with Dhaka emerging as a dominant hotspot surrounded by contrasting districts.

**Fig 1.**
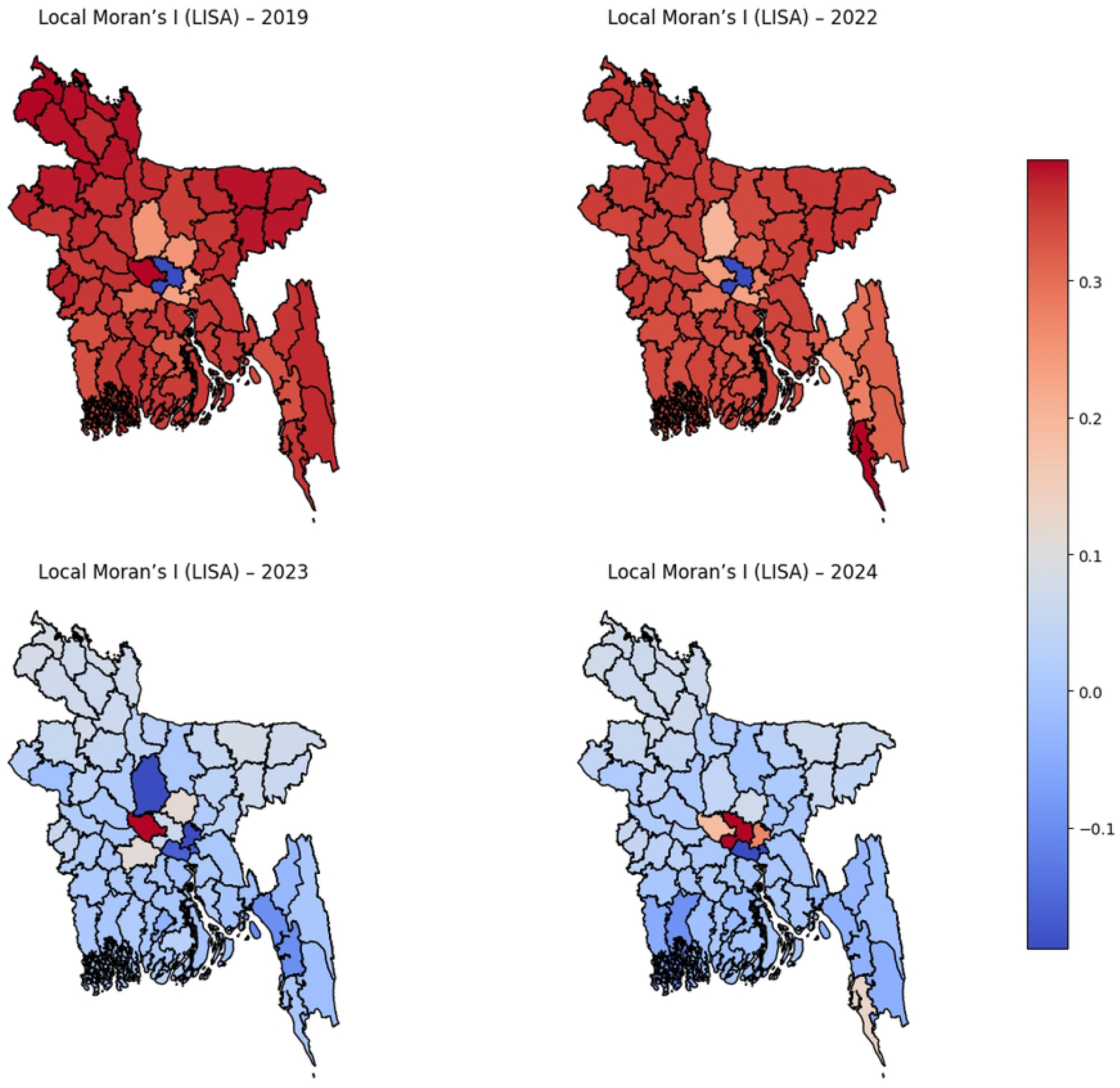
Local Moran’s I (LISA) cluster maps of district-level dengue incidence in Bangladesh, 2019–2024. The map illustrate significant high–high (red) and low–low (blue) clusters, along with high–low and low–high outliers, showing temporal shifts in spatial dependency across districts.

#### Temporal evolution and lagged effect dependency

The temporal evolution of dengue incidence in Bangladesh exhibits a pronounced seasonal pattern and an increasing long-term trend (Fig 2). Seasonal decomposition of the national time series (2000–2024) revealed that transmission consistently begins to rise in June, peaks between August and September, and declines by November. This seasonality aligns with the onset of the monsoon rains, which creates favourable breeding habitats for *Aedes* mosquitoes [47]. The trend component highlights a gradual increase in incidence from 2010 onwards, followed by a sharp escalation after 2015. This shift corresponds with the emergence of repeated large-scale epidemics, reflecting both intensified climatic variability and rapid urbanisation [48].

**Fig 2.**
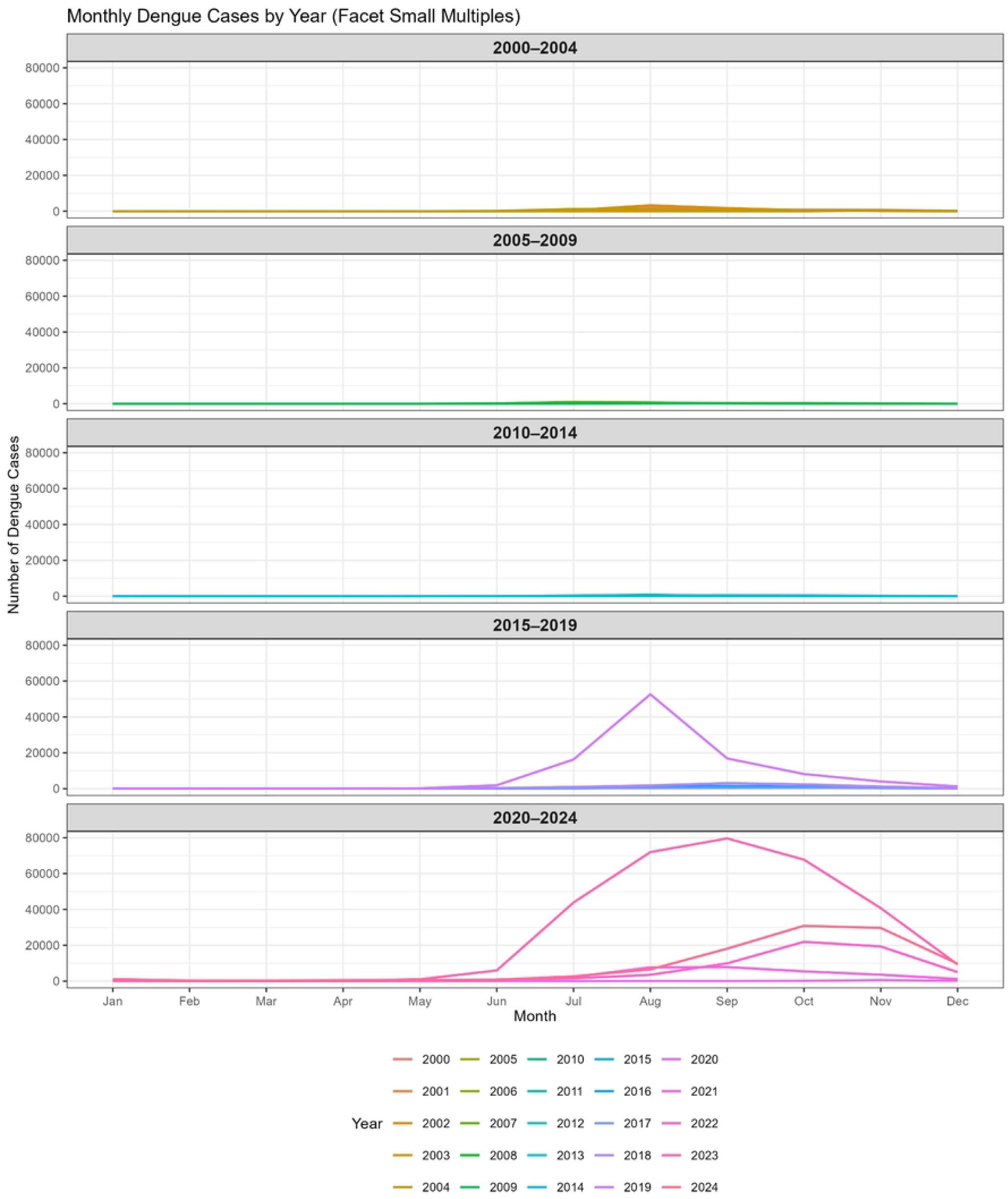
Seasonal decomposition of national dengue incidence in Bangladesh, 2000–2024. The faceted line plots show monthly dengue cases in Bangladesh across five periods: 2000–2004, 2005–2009, 2010–2014, 2015–2019, and 2020–2024. Each coloured line represents an individual year within the respective period. The y-axis indicates the number of reported dengue cases, while the x-axis represents months. The plots reveal minimal reported cases before 2010, a sharp rise with a major outbreak in 2019, and a substantial surge from 2020 to 2024, with cases peaking during the monsoon months (July–September).

The figure illustrates the seasonal, trend, and residual components of the dengue time series, showing consistent annual peaks during the monsoon period and a long-term upward trend. Boxplot analysis further confirmed that dengue incidence is concentrated during the monsoon months, with extreme outliers corresponding to major epidemic years (Fig 3).

**Fig 3.**
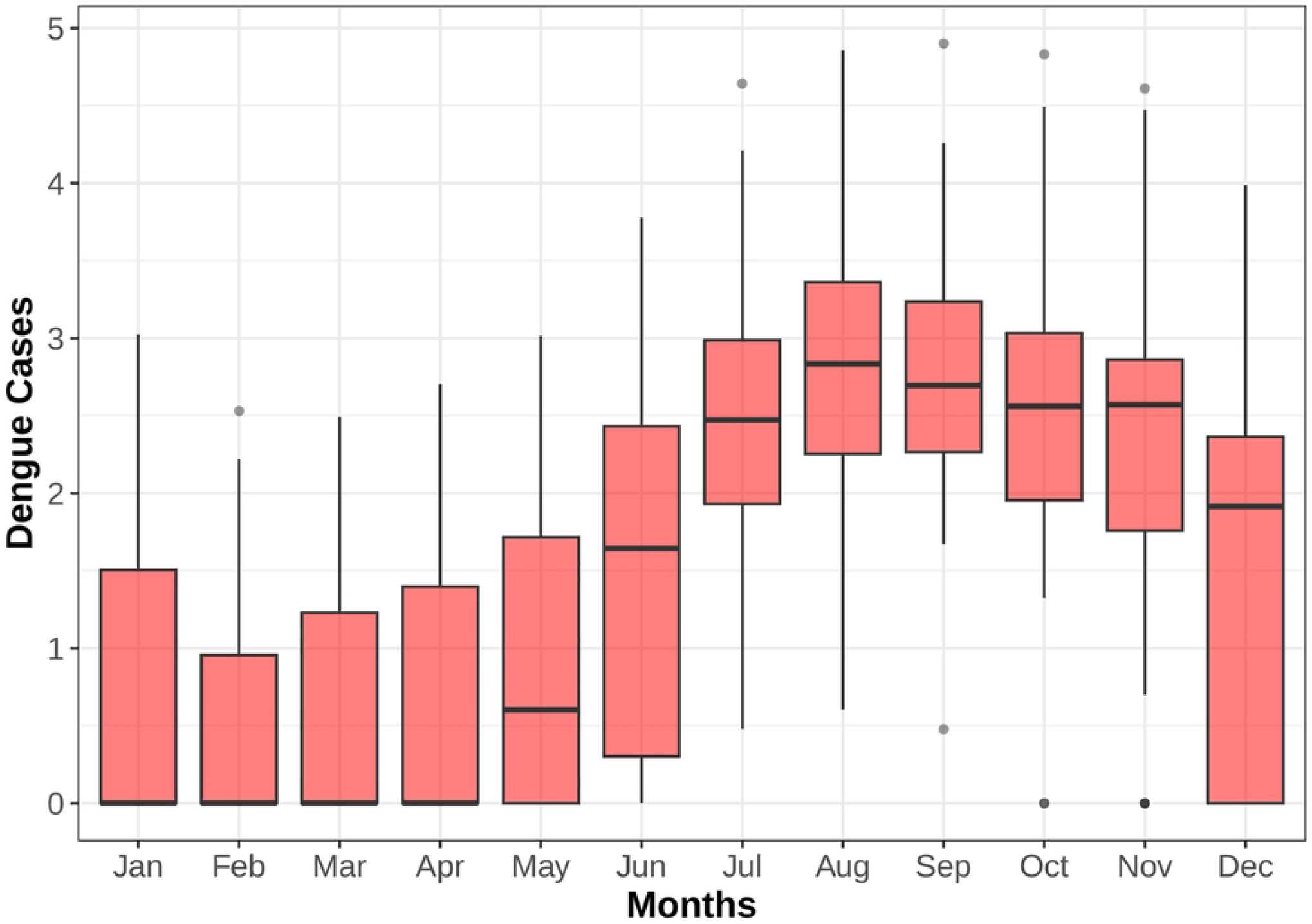
Boxplot of monthly dengue incidence in Bangladesh, 2000–2024. The boxplot displays the monthly distribution of dengue cases in Bangladesh from 2000 to 2024. The plot highlights the pronounced seasonal rise in dengue cases from June to September, peaking during the monsoon months, with lower incidence in the winter and early spring. This figure shows the concentration of cases during the monsoon months, with extreme values reflecting epidemic years and substantial inter-annual variability. The observed lagged dependency indicates that case counts peak approximately 4–8 weeks after the onset of rainfall, consistent with the extrinsic incubation period of the dengue virus and the life cycle of *Aedes* vectors [49,50]. The persistence of elevated incidence into October to November in recent years suggests a lengthening of the transmission season, likely linked to extended rainfall and higher post-monsoon temperatures. Such findings emphasise that dengue in Bangladesh is shaped not only by strong seasonal forcing but also by lagged climatic influences, underscoring the need for distributed lag and non-linear modelling approaches [51,52] to better capture the delayed impact of environmental drivers on dengue dynamics (Fig 4).

**Fig 4.**
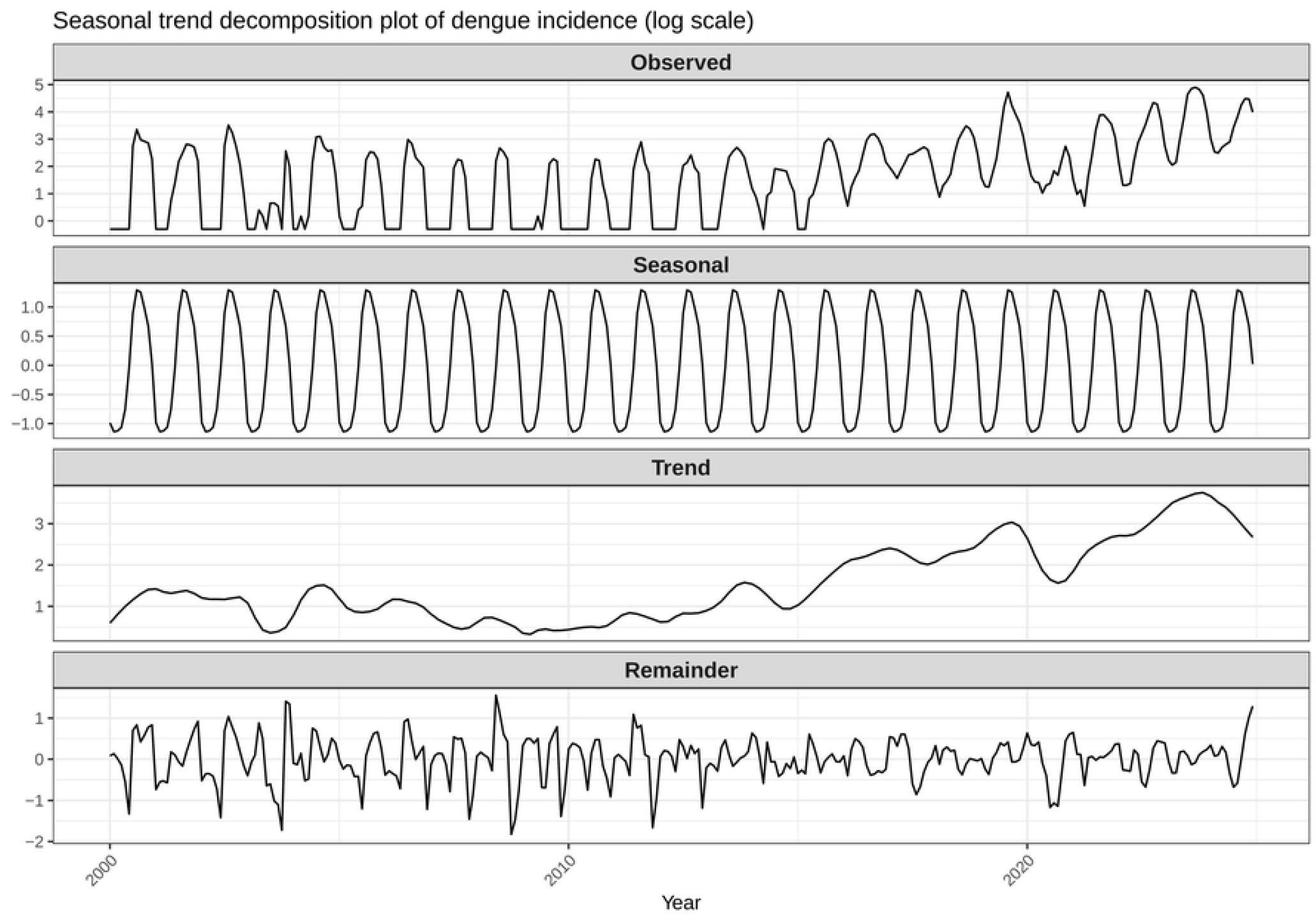
Lagged dependency between rainfall onset and dengue incidence in Bangladesh, 2000– 2024. This figure shows the seasonal-trend decomposition of dengue incidence in Bangladesh (2000–2024) on a logarithmic scale. The *Observed* panel represents the original time series of dengue cases. The *Seasonal* component captures the recurring yearly cycle of dengue incidence. The *Trend* component illustrates the long-term upward trajectory in dengue burden. The *Remainder* reflects irregular fluctuations and unexplained variability not accounted for by the seasonal or trend components.

#### Explanatory Temporal Analysis and Model identification

Stationarity of the dengue time series was confirmed using the Augmented Dickey–Fuller (ADF) test (ADF statistic = –8.41, p < 0.001), rejecting the null hypothesis of a unit root and validating the suitability of SARIMA modelling. Preliminary model identification was guided by inspection of the autocorrelation (ACF) and partial autocorrelation (PACF) functions (S7 Fig). The ACF displayed a damped sinusoidal decay with significant spikes at lags 1 to 3 and a seasonal peak at lag 12, suggesting short-term moving average and seasonal MA components. The PACF showed a sharp cut-off after lag 1, consistent with a first-order autoregressive term.

These diagnostic features motivated the specification of candidate SARIMA models with low-order AR and MA terms in combination with seasonal parameters. Model selection was refined by comparing alternative specifications using the Akaike Information Criterion (AIC). Among the competing models, SARIMA(1,0,2)(1,2,2)12 achieved the lowest AIC (4967.46), indicating superior fit relative to other candidates (S1 Table). Residual diagnostics further supported the adequacy of the selected model. The standardized residual series showed no obvious structure apart from isolated spikes, the correlogram indicated no significant autocorrelation, and Q–Q plots revealed approximate adherence to normality (S8 Fig). The Ljung–Box test confirmed that residuals were consistent with white noise. Collectively, these results demonstrate that the SARIMA(1,0,2)(1,2,2)12 model provides a robust characterisation of the temporal dynamics of dengue incidence in Bangladesh.

#### Climatic and demographic drivers of dengue incidence

Exploratory analysis indicates that climatic variability—rainfall, temperature, and humidity strongly modulate dengue incidence in Bangladesh (Fig 5).

**Fig 5.**
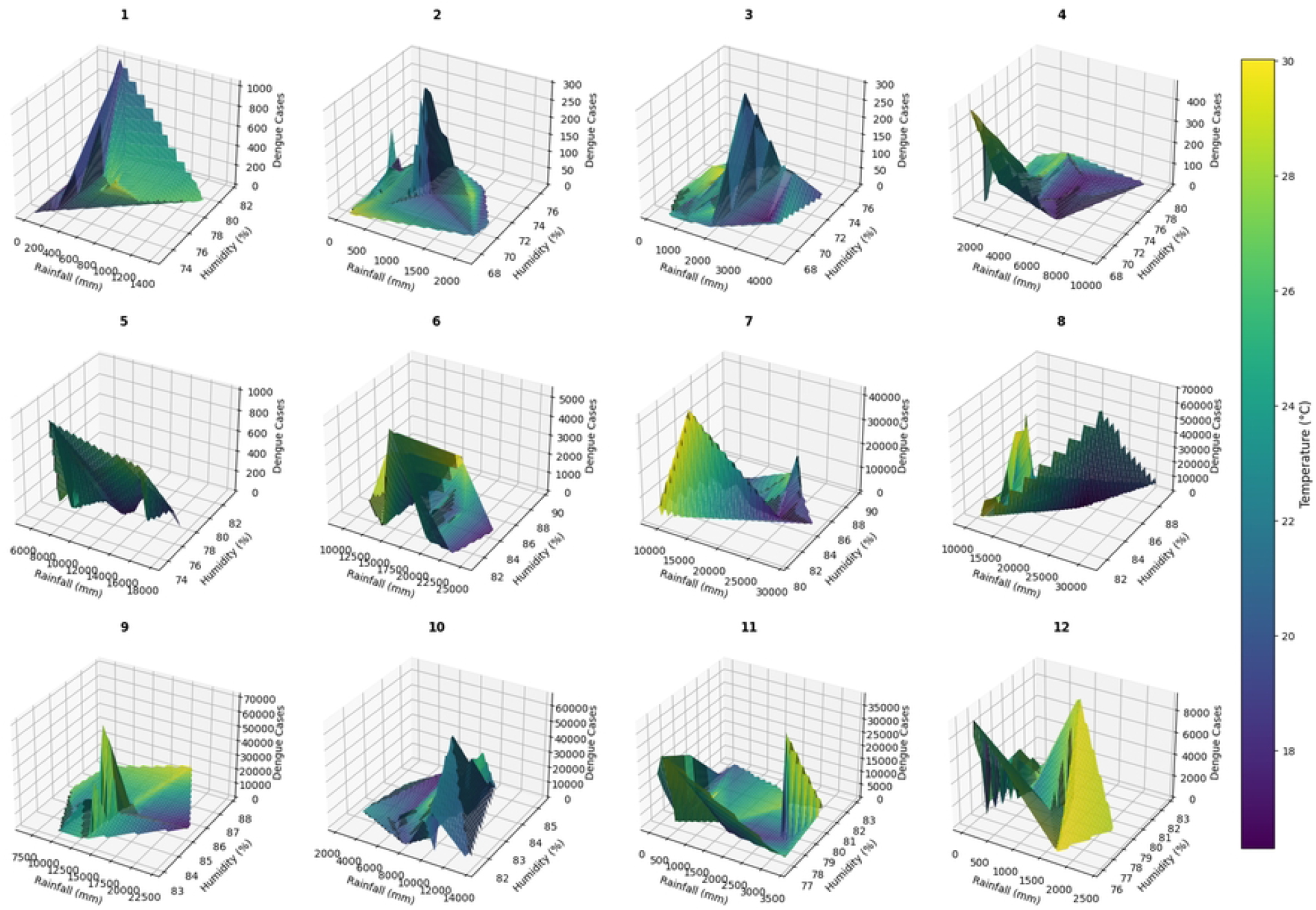
Surface plots showing the interaction of climate variables with dengue incidence across Bangladesh. In this figure, the colour gradient represents average monthly temperature (°C), ranging from cooler values (purple/blue) to warmer values (yellow/green). The x-axis indicates rainfall (mm), the y-axis shows relative humidity (%), and the z-axis represents reported dengue cases. Each numbered panel (1–12) corresponds to a calendar month, illustrating the combined influence of climatic drivers on dengue incidence. These plots of dengue cases against rainfall and humidity (with temperature as the colour scale) show pronounced seasonal structure: higher rainfall coupled with elevated humidity, within suitable temperature ranges, aligns with peaks in incidence, while less favourable combinations correspond to markedly lower case counts.

Inter-annual differences within the same month further illustrate sensitivity to year-to-year fluctuations in these climatic drivers. Demographic context also shapes transmission (Fig 6).

**Fig 6.**
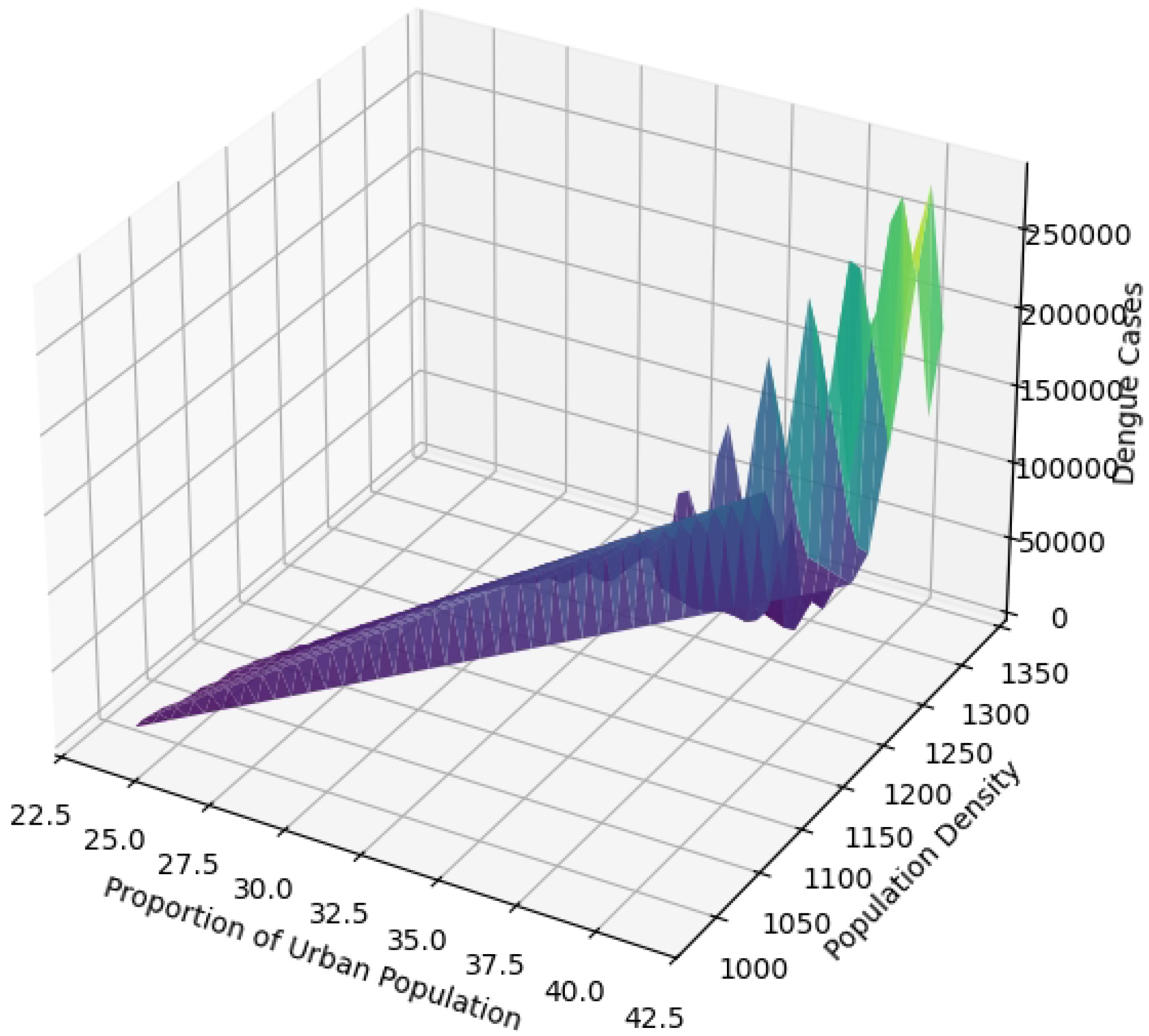
Surface plot of demographic drivers and dengue incidence in Bangladesh. The x-axis represents the proportion of the urban population (%), the y-axis indicates population density (persons/km^2^), and the z-axis shows reported dengue cases. The 3D surface plot reveals a strong positive association between demographic factors and dengue incidence. This figure shows Dengue cases increase sharply with higher population density, particularly when combined with a greater proportion of the urban population. The elevated peaks in the surface suggest that densely populated and highly urbanised districts are at substantially greater risk of large dengue outbreaks. This highlights the critical role of urbanisation and population clustering in shaping dengue transmission dynamics in Bangladesh, supporting the hypothesis that demographic pressure amplifies disease burden.

Because regression models were used, we assessed multicollinearity among covariates. The correlation heat map shows moderate associations among climatic variables and a strong correlation between the two demographic indicators (S9 Fig). Variance Inflation Factors (VIFs) confirmed the absence of severe multicollinearity for the explanatory variables (all VIF < 5; intercept not interpreted), supporting inclusion of all selected covariates with cautious interpretation of the closely related demographic pair (S2 Table).

#### Overdispersion and zero-inflation in dengue incidence data

Overdispersion is a common feature of disease count data [53]. In the context of dengue, the negative binomial model can adequately account for extra-Poisson variation [54]. In this dataset, the variance of dengue cases (8.45 × 10^6^) was substantially larger than the mean (2222.13), and a Poisson GLM yielded a dispersion statistic of 38,368.46, confirming strong overdispersion. To address this, we fitted a negative binomial model using maximum likelihood estimation implemented in Python (*statsmodels* library, Seabold & Perktold, 2010). The model provided a significantly better fit than the Poisson specification (LR statistic = 31,362,609.98, p < 0.001), supporting the negative binomial distribution as a more suitable framework for characterising dengue incidence. The negative binomial can also be interpreted as a compound Poisson distribution with a logarithmically distributed count per group [56], providing additional theoretical justification for its application in overdispersed infectious disease data. In addition, we employed Poisson mixture models within a SARIMA–Bayesian framework. This hybrid formulation offers a flexible alternative to the negative binomial by addressing extra-Poisson variation while enhancing forecasting reliability.

To further investigate excess zeros in the dengue dataset, we fitted both Zero-Inflated Poisson (ZIP) and Zero-Inflated Negative Binomial (ZINB) models using maximum likelihood estimation in Python (statsmodels library). However, both models exhibited convergence difficulties, with unstable coefficient estimates and non-significant inflation terms. For example, the Zero-Inflated Poisson (ZIP) model failed to converge (Log-Likelihood = –1,301,500, pseudo R^2^ = 0.05), and the Zero-Inflated Negative Binomial (ZINB) model similarly produced unreliable parameter estimates with undefined standard errors. Model comparison based on information criteria further indicated no improvement over the standard Negative Binomial model (Poisson AIC = 2,783,860; NB AIC = 4946; ZIP AIC = 2,662,999; ZINB AIC = not defined). These results suggest that zero-inflated formulations did not provide a better description of the data compared to the Negative Binomial specification, reinforcing the choice of NB-based and hybrid SARIMA–Bayesian models as more appropriate frameworks for modelling dengue incidence.

### Model developments

We employed a unified Bayesian hierarchical framework spanning both statistical and hybrid models. Classical formulations combined SARIMA with Poisson and Negative Binomial while Bayesian extensions were also applied to machine-learning hybrids (SARIMA–XGBoost, SARIMA–SVM, SARIMA–LSTM), enabling uncertainty quantification alongside predictive performance. The following section outlines the core components of these candidate models.

#### Poisson regression

Poisson regression provides a fundamental modelling framework for analysing count data, where the number of events in a fixed time interval is assumed to follow a Poisson distribution. The Poisson regression model represents the baseline approach for modelling count data, assuming equidispersion, i.e., the variance equals the mean of the response [57]. Let *Yt*denote the number of dengue cases at time t. The model is specified as:

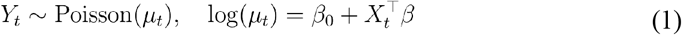

Here, *μ*_*t*_ is the expected number of cases and *X*_*t*_ denotes the vector of climatic and demographic predictors. Although widely used in epidemiology, its limitation lies in the strict equidispersion assumption, which is often violated in dengue surveillance data due to overdispersion.

The probability mass function is given by:

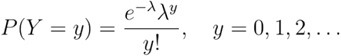

Where the mean and variance of the random variable are 𝔼 [*Y*] = *λ* and Var(*Y*)= *λ* respectively.

#### Negative binomial regression

The negative binomial formulation is employed to address overdispersion, with the parameter serving as a general control for this property [58]. Basically, negative binomial model extends Poisson regression to accommodate overdispersion by introducing a dispersion parameter r [58,59]. Its probability mass function is:

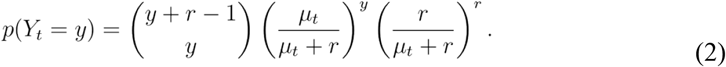

Where the mean and variance of the random variable are 𝔼 [*Y*_*t*_] = *μ*_*t*_ and 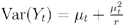.

As *r* → ∞, the distribution converges to Poisson.

#### Seasonal ARIMA (SARIMA)

To capture temporal autocorrelation and seasonality in dengue incidence, we adopt a Seasonal ARIMA (SARIMA) model [60]. The general form is:

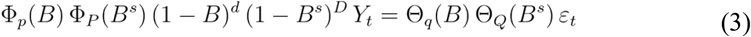

where B is the backshift operator, s denotes seasonality (12 months), and ℰ_*t*_ is white noise. This structure accounts for both short-term shocks and seasonal cycles in dengue cases [60].

#### Extreme Gradient Boosting (XGBoost)

XGBoost is an ensemble tree-based learning algorithm optimised for both speed and performance [61]. It employs gradient boosting, where regression trees are sequentially constructed to minimise a regularised objective:

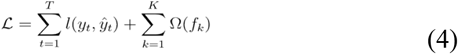

Where,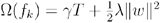

In our study, XGBoost was applied to capture nonlinear relationships between climatic and demographic covariates and dengue incidence and subsequently integrated with SARIMA residuals within a Bayesian framework.

#### Support Vector Machine (SVM)

Support Vector Regression (SVR) estimates a function *f*(*x*) that deviates from targets by no more than a threshold ϵ, while maintaining flatness [62,63]. The optimisation problem is:

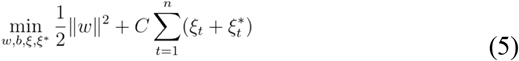

subject to margin constraints. Kernel functions enable nonlinear dependencies to be captured.

#### Long Short-Term Memory (LSTM)

The Long Short-Term Memory (LSTM) network is a recurrent neural network designed to overcome vanishing gradient problems by incorporating gated memory units [64,65]. At each time step t, information flow is regulated through forget, input, and output gates. The standard equations for forgetting, input, and output gates are:

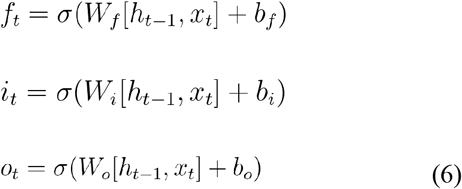

These gating mechanisms help the network retain and discard temporal information appropriately.

### Bayesian Statistical Mixture and Hybrid Machine Learning Modelling Framework

We evaluated a suite of statistical mixture and machine learning hybrid models of varying complexity, ranging from classical count regression to Bayesian SARIMA-based and deep learning extensions (Table 2). The structure and specification of each model are outlined in the following subsections.

**Table 2.** Model performance comparison based on leave-one-out information criterion (looic) for Bayesian SARIMA–mixture and hybrid models.

| S/N | Model | looic |
| --- | --- | --- |
| (A) | Bayesian SARIMA–Poisson mixture model | 131650.6842 |
| (B) | Bayesian SARIMA–NB mixture model | 2928.63487 |
| (C) | Bayesian SARIMA–XGBoost hybrid model | 1880.43858 |
| (D) | Bayesian SARIMA–SVM hybrid model | 5458.50238 |
| (E) | Bayesian SARIMA–LSTM hybrid model | 3303.018024 |

#### Bayesian SARIMA–Poisson mixture modelling Framework

We developed a Bayesian hierarchical Poisson–SARIMA model to account for temporal dynamics in dengue incidence. The observed counts *Y*_*t*_ were modelled as a Poisson process:

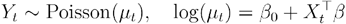

Where, *μ*_*t*_ denotes the mean case count at time t, *X*_*t*_ represents climatic and demographic covariates, and *η*_*t*_ captures latent temporal effects through SARIMA dynamics. The SARIMA component followed a (1,0,2) (1,2,2)12 specification to capture seasonality and autocorrelation:

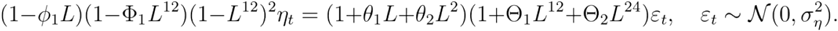

Priors were specified as weakly informative distributions to regularise estimation while avoiding undue influence on posterior inference. The regression coefficients (*β*_*0*_, *β*_*j*_)were assigned normal priors with mean zero and variance 10^2^. The SARIMA parameters, (*ϕ, θ*, Φ, Θ) were each given standard normal priors 𝒩(0,1), reflecting mild regularisation around zero. For the variance terms, an Inverse-Gamma prior with shape parameter 2 and scale parameter 0.5 was adopted, a common choice for modelling uncertainty in error variance under weak prior knowledge. This formulation enables joint modelling of climate–demographic covariates and temporal autocorrelation within a unified Bayesian framework, while quantifying uncertainty in both parameters and predictions.

#### Bayesian SARIMA–Negative Binomial mixture modelling Framework

To accommodate overdispersion in dengue incidence counts, we extended the Poisson formulation to a Negative Binomial regression framework with a SARIMA (1,0,2)(1,2,2)12 temporal component. At the likelihood level, observed counts *Y*_*t*_ at time t follow a Negative Binomial distribution:

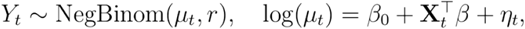

where *μ*_*t*_ is the expected mean, r is the dispersion parameter, *X*_*t*_ denotes climatic and demographic covariates, and *η*_*t*_ captures latent temporal dependency. At the process level, the latent component *η*_*t*_ follows a SARIMA (1,0,2) (1,2,2)12 specification:

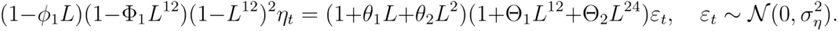

At the prior layer, regression coefficients were assigned weakly informative Gaussian priors, (*β*_*0*_, *β*_*j*_) were assigned normal priors with mean zero and variance 10^2^. The SARIMA parameters (*ϕ, θ*, Φ, Θ), were each given standard normal priors 𝒩(0,1), and for the variance terms, an Inverse-Gamma prior with shape parameter 2 and scale parameter 0.5 was considered. The overdispersion parameter was assigned a Gamma prior, *r*. ~ Gamma(2,0.1) This hierarchical specification allowed us to model both overdispersion and temporal autocorrelation, while quantifying parameter and predictive uncertainty within a Bayesian framework.

#### Bayesian SARIMA–XGBoost hybrid Modelling Framework

To jointly capture linear temporal dynamics and nonlinear covariate effects, we developed a Bayesian SARIMA–XGBoost hybrid model. The observed dengue counts *Y*_*t*_ were assumed to follow a count distribution, with the mean parameter expressed as a combination of SARIMA-based temporal structure and nonlinear contributions from XGBoost:

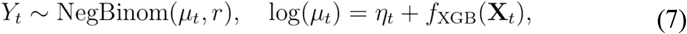

Where, *η*_*t*_ follows a SARIMA (1,0,2) (1,2,2)12 specification to capture seasonality and autocorrelation, and *f*_*XGB*_(*X*_*t*_) represents nonlinear effects estimated via an ensemble of regression trees. The Bayesian extension achieved by placing priors on SARIMA parameters (*ϕ, θ*, Φ, Θ) were each given standard normal priors 𝒩(0,1), XGBoost weights (*λ*_m_ were assumed to follow a normal prior 𝒩(0, τ^2^, with the scaling parameter *T* specified as a Half-Cauchy distribution with location 0 and scale 1. Variance terms (*σ*^2^) were modelled using an Inverse-Gamma prior with shape 2 and scale 0.5. To account for overdispersion, the dispersion parameter (*r*)was assigned a Gamma prior with shape 2 and rate 0.1.

This framework allowed both SARIMA and XGBoost to contribute simultaneously to the mean structure, with Bayesian inference providing parameter regularisation and predictive uncertainty quantification.

#### Bayesian SARIMA–SVM hybrid Modelling Framework

We developed a Bayesian SARIMA–SVM hybrid model to capture both linear temporal dynamics and nonlinear covariate effects in dengue incidence. The observed counts *Y*_*t*_ were modelled as a Negative Binomial to address overdispersion process with mean *μ*_*t*_.

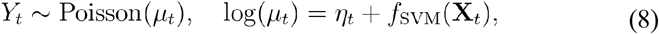

Where, *η*_*t*_ follows a SARIMA (1,0,2) (1,2,2)12 process accounting for seasonality and autocorrelation, *f*_*SVM*_ (*X*_*t*_) and captures nonlinear contributions via a Support Vector Machine kernel expansion. The SARIMA component evolves as

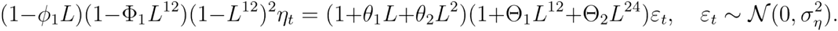

The SVM component is represented as, 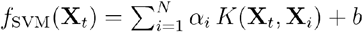, with kernel function *K*(·, ·), support vector weights α_*i*_ and intercept b. Within the Bayesian framework, weakly informative priors were assigned: regression and SARIMA parameters followed Gaussian priors, noise variances were given Inverse-Gamma distributions, and SVM coefficients were treated as random variables with Gaussian priors and Half-Cauchy hyperpriors for scale. This formulation allows the model to combine SARIMA’s strengths in handling seasonality and autocorrelation with SVM’s ability to capture nonlinear structures, while Bayesian inference provides coherent uncertainty quantification.

#### Bayesian SARIMA–LSTM Hybrid Modelling Framework

We developed a Bayesian SARIMA–LSTM hybrid model to capture both linear seasonal patterns and nonlinear temporal dependencies in dengue incidence. The observed counts were modelled as

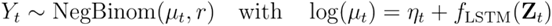

where *η*_*t*_ follows a SARIMA (1,0,2)(1,2,2)12 specification to account for seasonality and autocorrelation,*f*_*LSTM*_ (*Z*_*t*_)and captures nonlinear contributions from a Bayesian LSTM network. Priors were set as weakly informative for regression and SARIMA parameters (*ϕ, θ*, Φ, Θ) were each given standard normal priors 𝒩(0,1), with variance terms following inverse-gamma distributions, and LSTM weights assigned Gaussian priors with Half-Cauchy hyperpriors. This unified Bayesian framework enables dynamic forecasting while quantifying parameter and predictive uncertainty.

## Results and discussion

### Model assessment

Model performance was assessed using the expected log pointwise predictive density (elpd), estimated through leave-one-out cross-validation (LOO-CV), which approximates out-of-sample predictive accuracy [66]. For reporting, the leave-one-out information criterion (looic = –2 × elpdloo) was applied, providing a deviance-based measure consistent with the conventional Bayesian deviance information criterion (DIC) framework [67]. The Leave-One-Out Information Criterion (looic) values for all candidate models are summarised in (Table 2). Among the tested models, the Bayesian SARIMA–XGBoost hybrid model (Model C) achieved the lowest looic (1880.44), indicating the strongest predictive performance. The Bayesian SARIMA–NB mixture model (Model B) and the Bayesian SARIMA–LSTM hybrid model (Model E) followed with competitive fits, recording looic values of 2928.63 and 3303.02, respectively. By contrast, the Bayesian SARIMA–SVM hybrid model (Model D) and the Bayesian SARIMA–Poisson mixture model (Model A) exhibited considerably higher looic values, reflecting poorer predictive accuracy. Overall, these results emphasise the superiority of hybrid formulations that integrate machine learning components—particularly XGBoost—within the Bayesian SARIMA framework for forecasting dengue incidence.

Lower looic values indicate better model fit. Among the candidates, the Bayesian SARIMA– XGBoost hybrid model (C) achieved the lowest looic, reflecting superior predictive performance compared with Poisson, negative binomial, SVM, and LSTM formulations.

### Climate and demographic risk factors identification

Although the Bayesian SARIMA–NB mixture model (Model B) and the Bayesian SARIMA– LSTM hybrid model (Model E) exhibited considerable explanatory capacity, the Bayesian SARIMA–XGBoost hybrid model (Model C) emerged as the best-fitting specification according to the leave-one-out information criterion (looic) (Table 2). This model achieved the lowest looic, thereby outperforming all competing formulations, and was consequently selected to identify the climatic and demographic risk factors of dengue incidence.

Within the Bayesian SARIMA–XGBoost hybrid framework, rainfall and the proportion of the urban population were identified as significant predictors of dengue incidence. Rainfall exerted a consistently positive effect across all significance levels, with narrow credible intervals (e.g., mean coefficient = 0.0033; 95% CrI: 0.0015–0.0050 at the 5% level), indicating that increased rainfall contributes to a heightened risk of dengue transmission. Among demographic factors, the proportion of the urban population proved most influential, demonstrating robustly positive effects at the 5% level (mean coefficient = 1.9019; 95% CrI: 0.1645–3.7088). This finding underscores the critical role of urbanisation as a key driver of transmission dynamics (Table 3).

**Table 3.** Parameter estimates for climate and demographic covariates in the Bayesian SARIMA–XGBoost hybrid model (Model C).

| Variable | Level of significance<br>(0.1%) |  |  | Level of significance<br>(1%) |  |  | Level of significance<br>(5 %) |  |  |
| --- | --- | --- | --- | --- | --- | --- | --- | --- | --- |
|  | Mean<br>Coeff | Credible Interval |  | Mean<br>Coeff | Credible Interval |  | Mean<br>Coeff | Credible<br>Interval |  |
|  |  | 0.05% | 99.95% |  | 0.50% | 99.50% |  | 2.50% | 97.50% |
| Temperature | -0.124 | -2.977 | 2.4102 | -0.0987 | -2.3408 | 2.3272 | -0.213 | -1.906 | 1.4968 |
| Total Rain | 0.003 | 0.0004 | 0.0062 | 0.0032 | 0.0008 | 0.0053 | 0.0033 | 0.0015 | 0.0050 |
| Humidity | 0.700 | -1.415 | 3.001 | 0.7092 | -0.8886 | 2.4186 | 0.6894 | -0.529 | 1.9722 |
| Population<br>Density | -0.093 | -0.262 | 0.0896 | -0.0931 | -0.2299 | 0.0363 | -0.089 | -0.19 | 0.0107 |
| Proportion of<br>Urban<br>Population | 1.931 | -1.265 | 4.6673 | 1.9220 | -0.4866 | 4.1672 | 1.9019 | 0.1645 | 3.7088 |

The table presents posterior mean coefficients and 95% credible intervals (CrI) for climatic and demographic predictors at three significance thresholds (0.1%, 1%, and 5%). Rainfall and the proportion of urban population were identified as significant positive predictors of dengue incidence, while temperature, humidity, and population density showed non-significant associations.

### Prediction for dengue epidemics and an early warning system

To evaluate out-of-sample predictive performance, four model validation criteria were applied: root mean square error (RMSE), mean absolute error (MAE), continuous ranked probability score (CRPS) [68], and coverage probability at the 95% nominal level (CVG) [69]. RMSE and Mean Absolute Error (MAE) assess model accuracy in terms of mean response, whereas CRPS and CVG evaluate the quality of probabilistic forecasts. Specifically, CRPS quantifies the difference between observed values and the full predictive distribution [68], while CVG identifies potential underfitting or overfitting by examining deviations from the nominal 95% coverage probability [69]. For RMSE, MAE, and CRPS, smaller values indicate superior predictive performance. All validation metrics were computed in Python using the *scikit-learn* package for RMSE and MAE [70], and the properscoring library for CRPS [71]. For out-of-sample forecasting, exogenous variables were incorporated indirectly through the XGBoost and Bayesian components. While the SARIMA model produced baseline forecasts using only past case data [72], residuals unexplained by SARIMA were modelled using climatic and demographic covariates. The final dengue case forecasts thus reflected both the temporal dynamics captured by SARIMA [72] and the exogenous effects estimated via XGBoost [61] and Bayesian regression [73].

Similar hybrid frameworks combining statistical time-series models with machine learning methods have been successfully applied in infectious disease forecasting, including dengue [74]. The probability of dengue cases exceeding a defined threshold h at month t was estimated using the complementary cumulative distribution of the predictive posterior samples obtained from the hybrid SARIMA–Bayesian–XGBoost model. This probability was computed by drawing empirical samples of *Y*_*t*_ through Markov Chain Monte Carlo (MCMC) simulation and applying the indicator function:

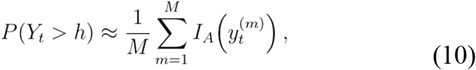

Where 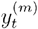 and M represents the *m*^*th*^ empirical sample of dengue cases at time t, and M denotes the number of empirical samples obtained from the MCMC algorithm. 𝕀(·) is an indicator function that the set A is the condition when the empirical sample from the posterior predictive distribution exceeds the threshold h (*i*.*e*.: *A* = {*x* ∈ ℕ_0_; *x* > *h*}) In this study, thresholds were defined to reflect epidemiologically meaningful alarm levels (e.g., the seasonal mean for that month, calculated from historical data (2000–2024)). This month-specific threshold accounts for dengue’s strong seasonal pattern in Bangladesh and reduces the likelihood of misclassifying expected seasonal peaks as outbreaks. By using the long-term monthly mean as the cut-off, only case counts surpassing the expected seasonal baseline were flagged, ensuring epidemiological relevance and avoiding false alarms [75,76]. Out-of-sample probability forecasts for the period 2022–2024 were generated using all candidate models. Model validation criteria were calculated based on the fitted models using data from 2016–2021, and performance was evaluated through RMSE, MAE, CRPS, CVG, and early warning system metrics of sensitivity and specificity [77,78] (Table 4).

**Table 4.**
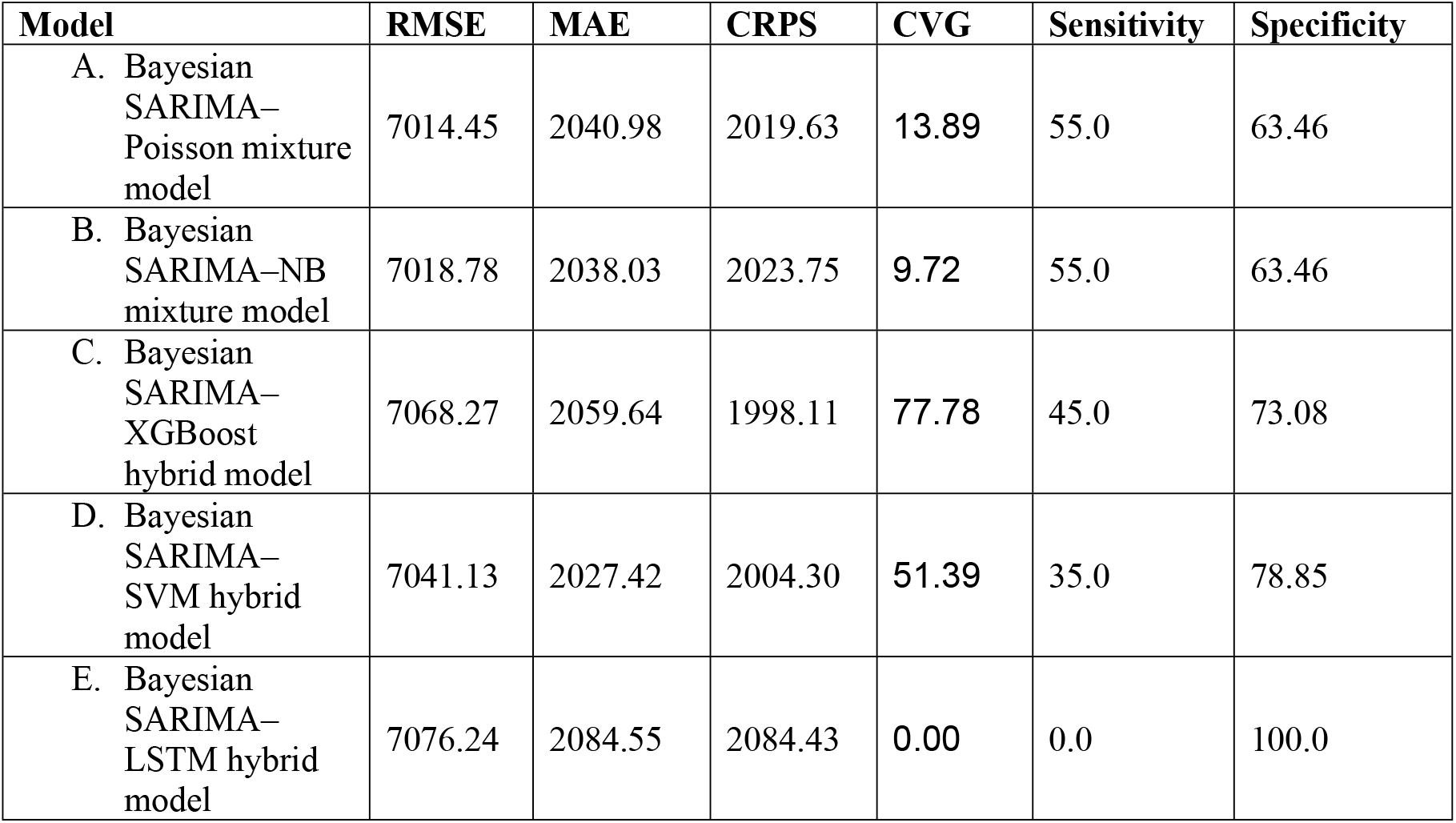
Model validation statistics and performance measures of early warning system derived from five models, A-E.

| Model | RMSE | MAE | CRPS | CVG | Sensitivity | Specificity |
| --- | --- | --- | --- | --- | --- | --- |
| A. Bayesian SARIMA–Poisson mixture model | 7014.45 | 2040.98 | 2019.63 | 13.89 | 55.0 | 63.46 |
| B. Bayesian SARIMA–NB mixture model | 7018.78 | 2038.03 | 2023.75 | 9.72 | 55.0 | 63.46 |
| C. Bayesian SARIMA–XGBoost hybrid model | 7068.27 | 2059.64 | 1998.11 | 77.78 | 45.0 | 73.08 |
| D. Bayesian SARIMA–SVM hybrid model | 7041.13 | 2027.42 | 2004.30 | 51.39 | 35.0 | 78.85 |
| E. Bayesian SARIMA–LSTM hybrid model | 7076.24 | 2084.55 | 2084.43 | 0.00 | 0.0 | 100.0 |

Among the competing models, the Bayesian SARIMA–XGBoost hybrid model (Model C) demonstrated the strongest performance, achieving the lowest CRPS (1998.11) and the highest CVG (77.78). This indicates a well-balanced trade-off between probabilistic accuracy and interval coverage [61]. In comparison, the Bayesian SARIMA–Poisson model (Model A) and the Bayesian SARIMA–NB mixture model (Model B) achieved slightly lower RMSE and MAE values; however, their CVG values were markedly lower (<15%), and their specificity was only moderate (63.46%). These results are consistent with earlier applications of Poisson and negative binomial time-series models in infectious disease surveillance [79]. The SARIMA–SVM hybrid model (Model D) exhibited high specificity (78.85%) but relatively low sensitivity (35.0%), reflecting the well-documented behaviour of support vector machines in imbalanced classification contexts [63]. By contrast, the SARIMA–LSTM hybrid model (Model E) yielded perfect specificity (100%) but failed to detect any outbreak months (0% sensitivity), underscoring the limitations of recurrent neural networks in capturing rare extreme events [64]. Taken together, these findings highlight the Bayesian SARIMA–XGBoost hybrid model (Model C) as the most effective specification, offering the best balance between predictive accuracy and outbreak detection. This makes Model C the most suitable candidate for national-level forecasting of monthly dengue cases.

The table reports root mean square error (RMSE), mean absolute error (MAE), continuous ranked probability score (CRPS), and coverage probability at the 95% level (CVG), alongside early warning system sensitivity and specificity. The Bayesian SARIMA–XGBoost hybrid model (C) achieved the lowest CRPS and highest CVG, indicating the most balanced predictive performance. In contrast, the SARIMA–LSTM hybrid model (E) demonstrated perfect specificity but failed to detect outbreak months (0% sensitivity).

### Out-of-Sample Forecast for 2025 and Implications for Early Warning

An out-of-sample probability forecast for the first eight months of 2025 (January–August) was generated using the best-fitting Bayesian SARIMA–XGBoost hybrid model to support early warning efforts. The analysis incorporated both historical observations (January 2022–August 2025) and forecasted values (January–August 2025). The model successfully reproduced the expected seasonal dynamics of dengue incidence. Low case numbers were projected for the early months (January–March), followed by a sharp increase during the monsoon period (May–July), with a peak of approximately 16,000 cases predicted in July before declining slightly in August (Fig. 7). Although the model tended to overestimate case magnitudes relative to observed data, the temporal alignment of outbreak peaks was well captured, a feature of particular importance for early warning system applications. Timely forecasts of this nature can enable public health authorities to pre-position resources, reinforce vector control strategies, and allocate clinical capacity ahead of peak transmission months, thereby mitigating the potential burden of large-scale outbreaks.

**Fig 7.**
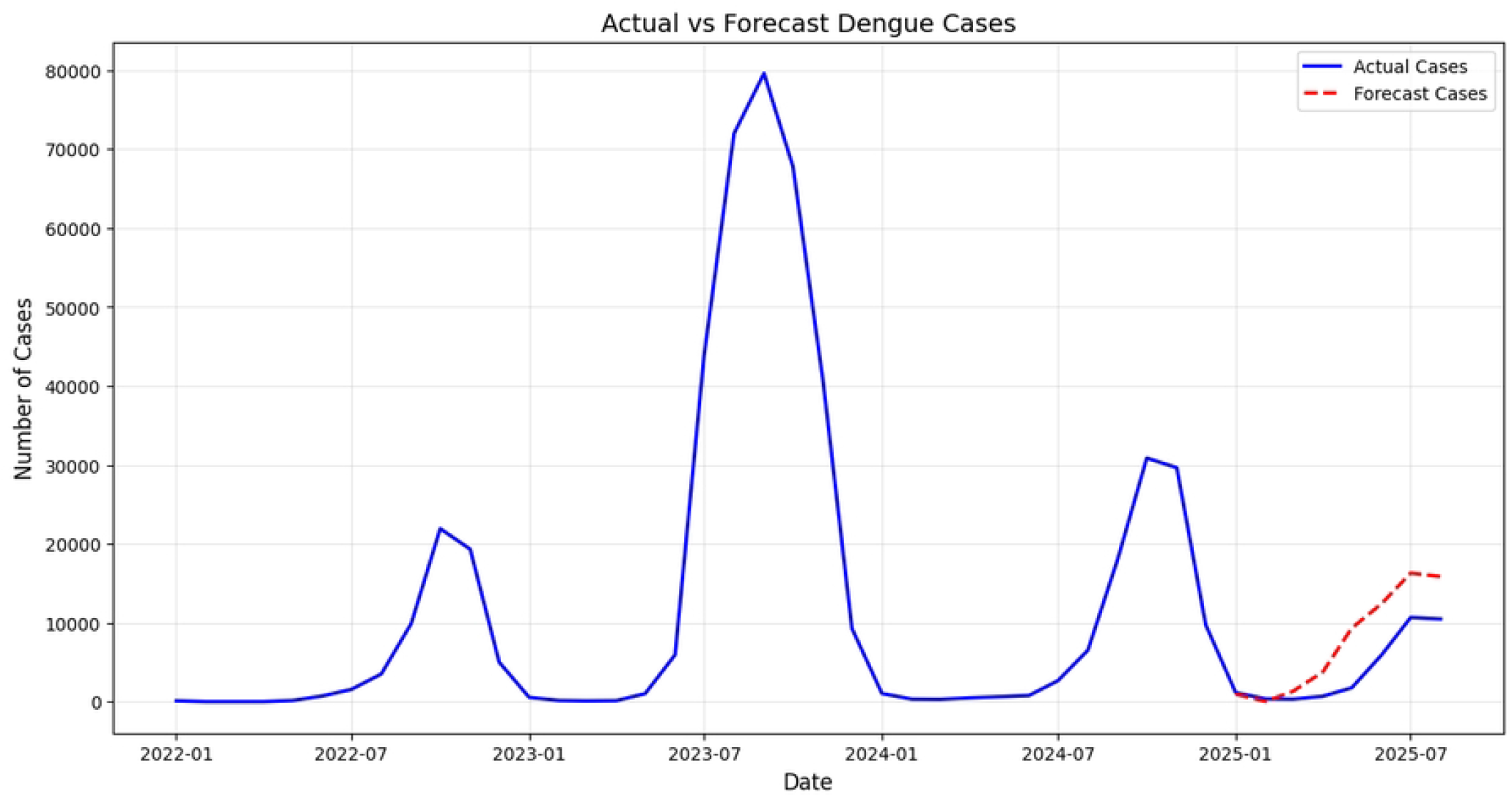
Comparison of observed dengue cases (January 2022–August 2025) and out-of-sample forecasts (January–August 2025). The line plot compares observed dengue cases (blue solid line) for the period 2022–August 2025 with model-forecasted cases (January to August, 2025). The x-axis represents time (monthly), and the y-axis indicates the number of reported dengue cases. The plot highlights the model’s ability to capture seasonal peaks, with forecasted values for 2025 aligning closely with the observed pattern and providing an early indication of potential case surges.

The forecasts are generated using the Bayesian SARIMA–XGBoost hybrid model. The results illustrate that the model captures the seasonal rise in incidence, with peak transmission anticipated during the monsoon months.

This study applied a comprehensive suite of Bayesian mixture and hybrid models to forecast dengue incidence in Bangladesh, using 25 years of national surveillance data enriched with climate and demographic covariates. Our analysis demonstrates that hybrid approaches integrating statistical and machine learning frameworks outperform classical regression-based models. In particular, the Bayesian SARIMA–XGBoost hybrid model consistently achieved the lowest looic and CRPS values, alongside the highest coverage probability (CVG), indicating superior predictive performance compared to Poisson, negative binomial, SVM, and LSTM alternatives.

Our findings corroborate recent work emphasising the limitations of classical Poisson and negative binomial models in the presence of over-dispersion and complex nonlinearities in dengue surveillance data [79,80]. While these models provided acceptable baseline fits, their predictive accuracy and coverage were markedly inferior to hybrid formulations. Previous studies in Bangladesh and other settings have similarly found that Poisson- and negative binomial-based time-series models capture broad seasonality but tend to underperform during peaks of epidemic outbreaks, where deviations from average patterns are not adequately represented [81]. The observed superiority of the XGBoost hybrid model aligns with regional and international studies where gradient boosting has outperformed both traditional time-series and deep learning methods for dengue forecasting [61].

Notably, the SARIMA–LSTM hybrid model failed to detect outbreak periods despite achieving perfect specificity, reflecting the challenges of recurrent neural networks in capturing rare extreme events with relatively small national-level datasets [64,82]. This finding is consistent with earlier studies in Brazil and Singapore, which highlighted the need for large training datasets and extensive hyperparameter tuning for deep learning models to deliver reliable epidemic forecasts [83,84]. Conversely, the SARIMA–SVM hybrid exhibited moderate accuracy but imbalanced sensitivity and specificity, consistent with the behaviour of support vector machines in imbalanced classification problems [63].

Beyond temporal dynamics, this study conducted spatial analysis using Global Moran’s I to assess district-level clustering of dengue incidence. No statistically significant global spatial autocorrelation was detected, suggesting that dengue incidence during 2019–2024 was not uniformly clustered across districts. Nevertheless, Local Moran’s I (LISA) cluster maps identified localised hotspots, highlighting that outbreak intensity remains spatially heterogeneous even when global clustering is absent. These findings underscore the importance of localised surveillance and control measures, consistent with other studies that have reported geographically constrained hotspots of dengue transmission [75,76].

Our analysis reinforces the pivotal role of rainfall and urbanisation as significant drivers of dengue transmission. Rainfall emerged as a consistent positive predictor across all significance levels, highlighting its contribution to creating breeding habitats for *Aedes* mosquitoes. Similarly, the proportion of the urban population showed a strong positive effect, underscoring how rapid urbanisation in Bangladesh amplifies the risk of dengue outbreaks. These findings are consistent with regional literature linking climatic variability and urban growth to heightened transmission. By contrast, temperature, humidity, and population density exhibited weaker or non-significant associations once rainfall and urbanisation were included, suggesting that their effects may be mediated through broader seasonal and demographic processes. A key contribution of this study is the development of a Bayesian temporal modelling framework that supports the implementation of an Early Warning System (EWS) for dengue outbreaks, linking upstream surveillance data to forecast production. By evaluating sensitivity and specificity, the study highlights the operational relevance of hybrid models. The Bayesian SARIMA–XGBoost hybrid provided the best balance between detecting true outbreaks and avoiding false alarms. Importantly, probability forecasts for January–August 2025 successfully captured the seasonal rise and peak timing of dengue, even though magnitudes were somewhat overestimated. In the context of public health, such conservative forecasts are preferable to underestimation, as the cost of missing an outbreak far outweighs the consequences of a false alert [79]. These results demonstrate the potential of hybrid Bayesian–machine learning models to serve as the backbone of an operational EWS, enabling health authorities to pre-position resources, strengthen vector control, and enhance clinical preparedness ahead of peak transmission months.

The principal strengths of this study include its long national time series (2000–2024), integration of climate and demographic covariates, and rigorous comparison of multiple Bayesian mixture and hybrid models. The unified Bayesian framework enabled uncertainty quantification and probabilistic forecasting, while the spatial analysis component provided insights into heterogeneities across districts. However, several limitations must be acknowledged. First, the spatial analysis was restricted to district-level data for only 2019 and 2022 to 2024, which may not fully capture longer-term geographic clustering. Second, surveillance data may suffer from underreporting or diagnostic inconsistencies, particularly in earlier years, which could influence model calibration. Third, while exogenous variables were incorporated indirectly through XGBoost and Bayesian components, real-time climate forecasts and entomological indices were not included, limiting operational applicability. Future research should prioritise finer spatiotemporal resolution, leveraging district-level data and Bayesian spatiotemporal frameworks such as INLA to capture geographic heterogeneity. Integration of real-time meteorological forecasts and entomological indicators could further enhance early warning capabilities. Hybrid extensions using ensemble learning or attention-based deep learning models (e.g., Transformers) also merit exploration for improving the capture of rare epidemic dynamics.

## Conclusion

This study demonstrates the utility of Bayesian hybrid modelling frameworks for forecasting dengue incidence in Bangladesh. Among the candidate models, the SARIMA–XGBoost hybrid achieved the most favourable balance of predictive accuracy and outbreak detection, outperforming classical Poisson, negative binomial, and other machine learning hybrids. Incorporating rainfall and urbanisation as key covariates significantly improved forecasts, while spatial analysis revealed the absence of global clustering but highlighted localised hotspots through LISA. By linking upstream surveillance data to probabilistic forecasts, the proposed Bayesian temporal framework establishes the foundation for an operational early warning system. Although case magnitudes were sometimes overestimated, the accurate capture of outbreak timing underscores the model’s value for anticipatory public health action. These forecasts can support timely mobilisation of resources, targeted vector control, and clinical preparedness.

Future research should focus on extending the framework to finer spatial scales, incorporating real-time climate forecasts and entomological data, and testing advanced ensemble or attention-based deep learning models. Strengthening collaborations with health authorities will be essential to ensure the translation of these methodological advances into actionable dengue early warning systems.

## Data Availability

Dengue case and mortality data were obtained from the Directorate General of Health Services (DGHS), Bangladesh (https://old.dghs.gov.bd/index.php/bd/home/5200-daily-dengue-status-report). District-level population density and urban proportion data are publicly available from the Bangladesh Bureau of Statistics (https://bbs.gov.bd/). Monthly climate variables (temperature, rainfall, and humidity) were acquired from the Bangladesh Meteorological Department (BMD) via their online request system (https://live6.bmd.gov.bd/) and require purchase. Data, together with the R and Python analysis code used to generate the results and selected figures, are provided in the Supporting Information and in the Zenodo repository (https://doi.org/10.5281/zenodo.17099754). Please note that BMD does not permit redistribution of the purchased raw climate data without explicit written permission.

## Declaration of Competing Interest

The authors declare that they have no known competing financial interests or personal relationships that could have appeared to influence the work reported in this paper.

## Acknowledgments

The acknowledge with gratitude the Directorate General of Health Services (DGHS), the Bangladesh Bureau of Statistics (BBS), and the Bangladesh Meteorological Department (BMD) for providing the data used in this study.

## Supporting information

**S1 Fig. Annual dengue cases and deaths in Bangladesh (2000–2024, log10 scale)**. This figure shows the yearly distribution of dengue cases and deaths reported nationally between 2000 and 2024. Both series are presented on a log10 scale to visualise long-term trends, highlight epidemic peaks, and reduce skewness caused by extreme outbreak years.

**S2 Fig. Division-wise distribution of total dengue cases in Bangladesh, 2019–2024**. This choropleth map shows the total reported dengue cases across the eight administrative divisions of Bangladesh during the last five years. Dhaka and Chattogram divisions reported the highest case burdens, while peripheral divisions such as Rangpur and Sylhet consistently showed lower incidence.

**S3 Fig. District-wise monthly distribution of dengue cases in Bangladesh, 2022**. This heatmap displays the spatial distribution of dengue cases across districts by month for 2022. High transmission was concentrated in central and southeastern districts, whereas northern and coastal areas experienced low incidence

**S4 Fig. District-wise monthly distribution of dengue cases in Bangladesh, 2023**. The figure illustrates monthly dengue case distribution across districts in 2023. Central districts, particularly Dhaka and surrounding areas, recorded disproportionately higher case numbers compared with many northern districts.

**S5 Fig. District-wise monthly distribution of dengue cases in Bangladesh, 2024**. This heatmap depicts dengue case distributions by district for 2024. Consistent hotspots are observed in Dhaka, Chattogram, and neighbouring districts, while many peripheral districts continued to report very low numbers

**S6 Fig. Annual distribution of dengue cases by district in Bangladesh, 2000–2024 (log**_**10**_ **scale)**. This boxplot shows the variation in annual dengue incidence across districts over 25 years, expressed on a log_10_ scale. Substantial heterogeneity is evident, with certain districts showing persistently high incidence while others remain at the lower end of the distribution

**S7 Fig. Autocorrelation function and partial autocorrelation function of monthly dengue incidence in Bangladesh (2000–2024)**. The left panel shows the autocorrelation function (ACF), while the right panel presents the partial autocorrelation function (PACF) of the monthly dengue time series. The plots illustrate significant autocorrelation at short lags and inform the selection of appropriate SARIMA model parameters.

**S8 Fig. Diagnostic plots of standardized residuals from the fitted SARIMA model**. The standardized residual time series (top left) shows no obvious structure apart from occasional spikes. The histogram with kernel density estimate (top right) and the Q–Q plot (bottom left) indicate departures from normality, with heavier tails. The correlogram (bottom right) suggests that residual autocorrelations lie mostly within the 95% confidence bounds, indicating adequate model fit.

**S9 Fig. Correlation structure of climatic and demographic covariates considered in dengue models**. The heatmap displays pairwise correlation coefficients among climatic (temperature, total rainfall, humidity) and demographic (population density, proportion of urban population) covariates.

**S1 Table. Comparison of SARIMA candidate models based on AIC**. The table presents alternative SARIMA specifications fitted to monthly dengue incidence in Bangladesh, 2000–2024. The model SARIMA(1,0,2)(1,2,2)12 achieved the lowest AIC value (4967.46), indicating the best overall fit among the candidate models.

**S2 Table. Variance Inflation Factor (VIF) values of climatic and demographic explanatory variables included in the regression models**. The table presents VIF estimates used to assess multicollinearity among covariates. All explanatory variables show acceptable VIF values (<5), indicating no serious multicollinearity issues.

## Notes

### Competing Interest Statement

The authors have declared no competing interest.

### Funding Statement

The author(s) received no specific funding for this work.

### Author Declarations

This study analysed publicly available, de-identified secondary data on dengue cases and climate variables. No human participants or identifiable personal information were involved. According to the policies of the Directorate General of Health Services (DGHS), Bangladesh Bureau of Statistics (BBS), and Bangladesh Meteorological Department (BMD), no institutional review board or ethics committee approval was required.

